# The KCNT1-related epilepsy study: design and methods of a fully-decentralized prospective natural history study in a rare disease

**DOI:** 10.1101/2025.11.11.25340032

**Authors:** Heather R. Adams, Viet Nguyen, Laurie Seltzer, Carolyn Dickinson, Courtney Aponte, Sarah Hubbard, Marco Rizzo, David R. Bearden

## Abstract

**Objective:** KCNT1-related epilepsy is an ultra-rare pediatric onset epileptic encephalopathy with a broad clinical phenotype ranging from, most commonly, severe infantile-onset epilepsy and global developmental delay to, less commonly, milder phenotypes including nocturnal seizures, autism spectrum disorder, and learning disability. We initiated the first-ever prospective natural history study to comprehensively clinically phenotype individuals impacted by this disorder.

**Methods:** The primary study aim was to characterize seizures in individuals with KCNT1-related epilepsy. Secondary and exploratory aims included characterization of the full spectrum of disease symptoms, understanding caregiver burden, and collection of blood and urine samples for biomarker exploration. All study activities were conducted remotely (e.g., home-based assessments, telehealth visits).

**Results:** 35 participants (n=20 males, 15 females) enrolled in this study. The average age at the baseline visit was 76.0 months old (s.d. = 75.5). This paper presents the study design and methods, presents several challenges that arose in its implementation, and discusses various solutions implemented in this medically complex population.

**Significance:** Future work will apply the lessons from the current study in the planning and design of clinical trials for KCNT1-related epilepsy and possibly other developmental and epileptic encephalopathies.

**Highlights:**

- KCNT1-related epilepsy is an ultra-rare pediatric onset epileptic encephalopathy with no disease-modifying therapy yet available.
- Because of its rarity, little is known about the phenotypic range and natural history of symptoms in affected individuals.
- To inform clinical trial planning, we initiated the first ever prospective, longitudinal natural history study of KCNT1-related epilepsy.
- The unique all-remote design of this study presented various challenges, opportunities, and learnings that will inform future studies.

## 1. Introduction

KCNT1-related epilepsy is an ultra-rare genetic disorder resulting from de novo or inherited pathogenic variants in the KCNT1 gene. KCNT1 encodes the “Slack” or Slo2.2 sodium-activated potassium channel, affecting neuronal excitability through its role in neuronal repolarization and hyperpolarization following action potentials. Incidence or prevalence are not well known. A recent study in Denmark reported a birth incidence of ≤1.12 per 100,000 live births, and an annual incidence rate of 0.04 per 100,000 persons [1]. A prior, two-year study of patients in the United Kingdom found slightly higher birth incidence of 0.26 and 0.55 cases per 100,000 live births in the first and second years of the study, respectively, leading the authors to estimate UK prevalence to be 0.11 per 100,000 persons across the entire study period [2]. However, estimated incidence and prevalence may increase as gene panel, exome, and genome testing become more commonplace in pediatric epilepsy, and research and awareness of this disorder grow. For example, patient advocacy efforts powered by social media and word-of-mouth have identified around 500 confirmed cases globally, half of which were identified in the United States [3].

The most common and severe clinical phenotypes of KCNT1-related epilepsy are Epilepsy of Infancy with Migrating Focal Seizures (EIMFS) and early-onset epileptic encephalopathies (EOEE). Both involve multiple, treatment-resistant daily seizures beginning in the first 6 months of life, acquired microcephaly, and profound intellectual and developmental delay [4–6]. Individuals with EIMFS or EOEE are also at high risk for sudden unexpected death in epilepsy [5, 7], A less severe phenotype is sleep-related hypermotor epilepsy (SHE), previously called autosomal dominant nocturnal frontal lobe epilepsy (ADNFLE). Seizure onset for individuals with SHE commonly begins in the first decade of life, and can be accompanied by: intellectual and developmental delay, autism spectrum disorder (mild to profound), executive dysfunction, psychiatric symptoms, and/or impaired sleep [6, 8].

Irrespective of phenotype, no available disease modifying therapies exist; care is supportive and focused on symptom management, particularly seizure control. However, there is little systematic knowledge regarding clinical phenotype, symptom progression, and clinical outcomes for affected individuals as a group, or within phenotype. Therefore, current management strategies are individually based, and further guidance is desirable regarding the collective patient experience. Additionally, with multiple drug development efforts in progress, including potential disease-modifying therapies, there is a need to establish valid and responsive clinical outcome measures that are relevant and meaningful for patients and families. It would also be helpful to identify biomarkers sensitive to disease state and to provide precise and responsive signals for efficacy and safety monitoring. The authors therefore established “A Natural History Study of Participants With Potassium Sodium-Activated Channel Subfamily T Member 1 (KCNT1)-Related Epilepsy (K1Te; clinicaltrials.gov registration: NCT04924153). The study goals were to characterize the natural history of KCNT1-related epilepsy and inform biomarker discovery to aid future clinical studies. The study’s primary endpoint was the seizure frequency and type experienced by affected individuals. Secondary and exploratory study objectives included measurement of various infant and child developmental and growth milestones, understanding the full spectrum of KCNT-1 related symptoms and their management, collection of various fluid samples for biomarker and genetic analyses, and assessment of adaptive function, quality of life, and caregiver burden.

## 2. Study Design

### 2.1 Methods - Overview

The K1Te Study was a nonrandomized, non-drug, 12-month prospective, longitudinal, fully-decentralized single-center study conducted by the University of Rochester Medical Center (URMC), Rochester, New York. Study activities were completed via in-home visits by contracted health care vendors, virtual appointments with study personnel, and questionnaires. The investigators, study sponsor team, and contract research organization (CRO) met weekly to discuss study progress.

#### 2.1.1 Recruitment strategy and advocacy group partnership

The KCNT1 Epilepsy Foundation, a national nonprofit organization with global outreach provided input on study design and heavily supported recruitment. IRB-approved recruitment materials were disseminated by the Foundation and to participants enrolled in the KCNT1/Migrating Partial Seizures of Infancy contact registry at URMC.

#### 2.1.2 Inclusion/Exclusion criteria

Target enrollment was 40 participants, based on the number of affected individuals within the contact registry and known through collaboration with the KCNT1 Epilepsy Foundation. Eligible participants were individuals from birth through 50 years of age with clinical signs and a confirmed genetic diagnosis of KCNT1-related epilepsy. Individuals were excluded if their mutation was a known benign variant. Other exclusion criteria were: HIV infection history, CNS tumors, malignancies or metastatic disease; current or past enrollment in an investigational gene therapy study, current enrollment in an interventional study of an investigational or approved drug, positive urine pregnancy test, and any condition that in the judgment of the PI (D.B.) would interfere with assessment of KCNT1-related epilepsy features and was not clearly related to that primary diagnosis. Any and all concomitant therapies including prescription and nonprescription medications, supplements, vaccines, etc., were allowed; their use was neither prescribed nor modified by the investigator.

#### 2.1.3 Visit Structure

Participants completed 6 study visits, which participants could choose to conduct remotely or on site. A Screening Visit preceded the Baseline/Visit 1 (within 30 days) to review inclusion/exclusion criteria and obtain informed consent (Parental Permission). Study activities included collection of medical history, demographics, history of seizures and anti-seizure treatment, completion of a seizure diary, use of a home-based wearable device (for seizure tracking), and identification of up to four participant-specific ‘most concerning symptoms’ tracked throughout the study using a visual analog scale. Study Visits 1-5 were performed at 3-month intervals. If participants withdrew prematurely (self- or investigator-directed), the study team attempted to conduct an end-of-study visit per the scheduled activities for Visit 5.

Before each study visit, the Coordinator (C.D.) contacted families to schedule study activities. The Coordinator and PI met with the participant and caregiver(s), in all cases one or both of the child’s parents, at the beginning of each study visit via telehealth (secure Zoom meeting). During this meeting, medication reconciliation and a physical and neurologic exam were conducted, and the Clinical Global Impression (CGI) scale (description below) was completed. Parents received education on diary compliance, and were asked to alert the study team to any adverse or reportable events including new symptoms, hospitalizations, or changes in medications or seizure type. Separately, the Vineland Adaptive Behavior Scales- Third Edition (Vineland-3) Comprehensive Interview was completed with a study Psychologist (C.A., H.R.A.) via telehealth [9].

A home-based phlebotomy service collected blood samples for blood chemistry and hematology, biomarkers, and DNA analysis. Other home health assessments at Visit 1 included: height, weight, and body mass index (BMI) calculation, vital signs, and 12-lead electrocardiogram (ECG). At Visits 1, 3, and 5, an overnight EEG and head circumference measurement was completed.

Parents maintained a daily seizure diary throughout study participation, and a one-week sleep diary at the time of each study visit. At Visits 1-5, parents also completed assessments of quality of life, sleep, mood, and behavior. Adults with a milder KCNT1-related epilepsy disorder phenotype and capacity to self-report on their function completed patient-reported outcome measures of mood, behavior, and the presence of psychotic symptoms.

### 2.2 Seizure outcome measures

#### 2.2.1 Electronic Seizure diary

The primary study endpoints were seizure frequency and type as recorded by parents in a daily electronic seizure diary. The diary captured the number, date, portion of day (in 6-hour blocks) of seizures, seizure type, and if rescue medication was used.

#### 2.2.2 Overnight Electroencephalogram (EEG)

A qualified vendor conducted overnight, home-based video EEG that was monitored remotely by an EEG technologist. Electrodes were applied using the standard International 10-20 system. The EEG unit was battery-operated. Data were saved to a storage card, then uploaded to a secure server. Parents pushed an event-button to mark the EEG if a clinical seizure was noted or suspected, and recorded the time and observations in an event log. Parents received extra supplies (e.g., tape) to re-secure exposed or loose electrodes, and a guide on positioning monitoring equipment (laptop, camera) and ensuring EEG recording equipment was secure and safe on their child. A 24/7 tech support contact phone number was provided, and the remote technologist contacted parents if problems were noted remotely, e.g., loss of laptop signal or a problem with an electrode. The EEG technologist returned to the home the following day to disconnect the EEG. Recordings were approximately 12 hours long, to capture awake, drowsy, and sleep states.

All EEGs were centrally read by a pediatric epileptologist on the study team (L.S.) who coded EEGs for seizure type, frequency, duration, background features including organization and interictal epileptiform discharges, and sleep architecture.

#### 2.2.3 Wearable device

A wrist-worn wearable device from a qualified vendor was available for up to 15 participants to capture exploratory endpoints. The device did not perform automated seizure detection, but captured physiologic and movement data (including possible seizures).

### 2.3 Adaptive Function

The Vineland Adaptive Behavior Scales, Third edition, Comprehensive Interview (Vineland-3) [9] is a proxy (parent) assessment of the ability to independently perform various age-expected skills. The Vineland-3 assesses functional domains of communication, daily living skills, socialization, and motor skills. Because participants’ chronological age was typically greater than developmental level, we estimated a composite developmental age score by calculating the mean age-equivalent (AE) score across all non-motor Vineland-3 subdomains. The mean AE then guided selection of other assessments (see Table 1). For example, a 10-year old child with a mean Vineland-3 AE=3 years would be assessed with measures suitable for a 3 year old child.

**Table 1.**
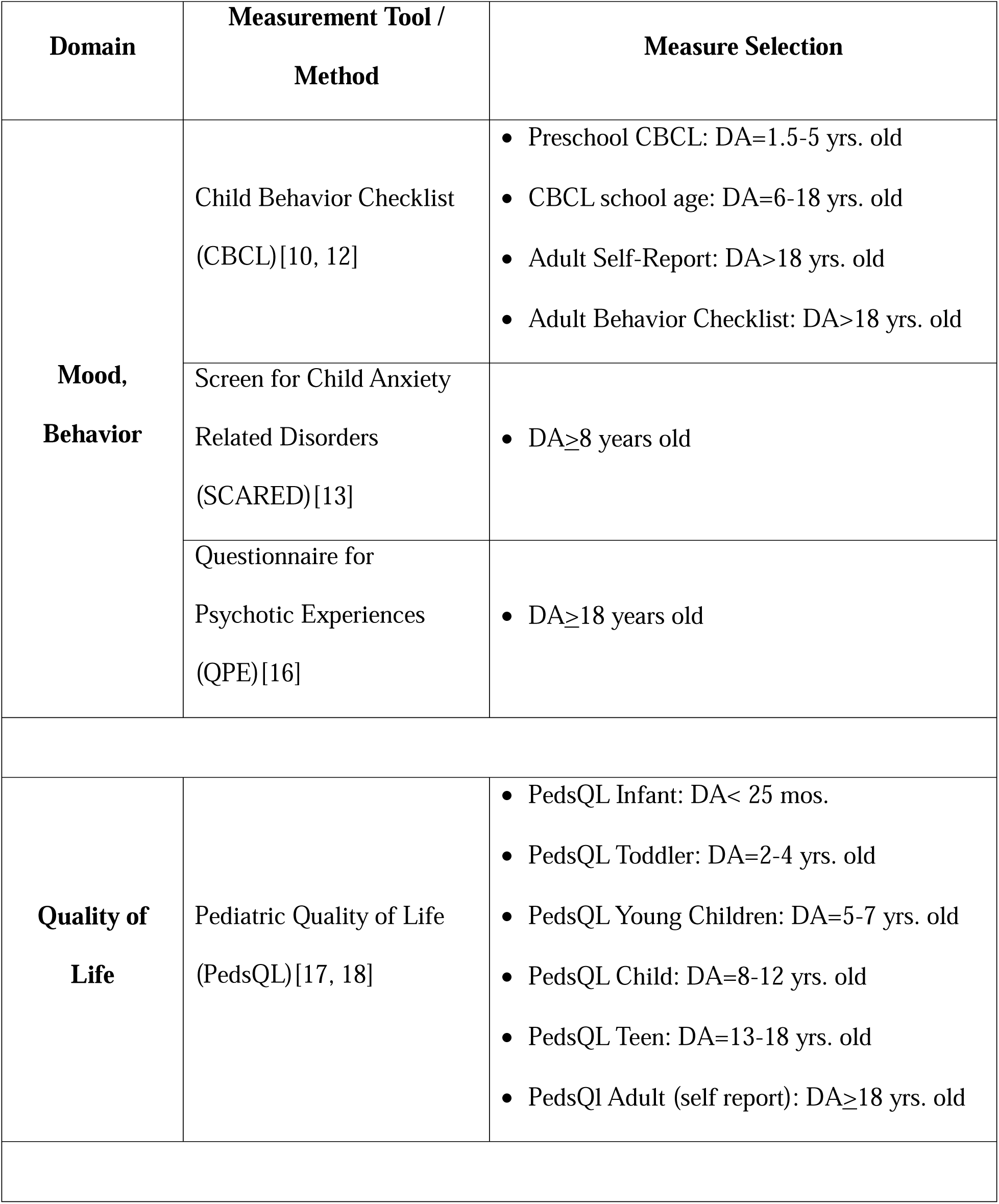
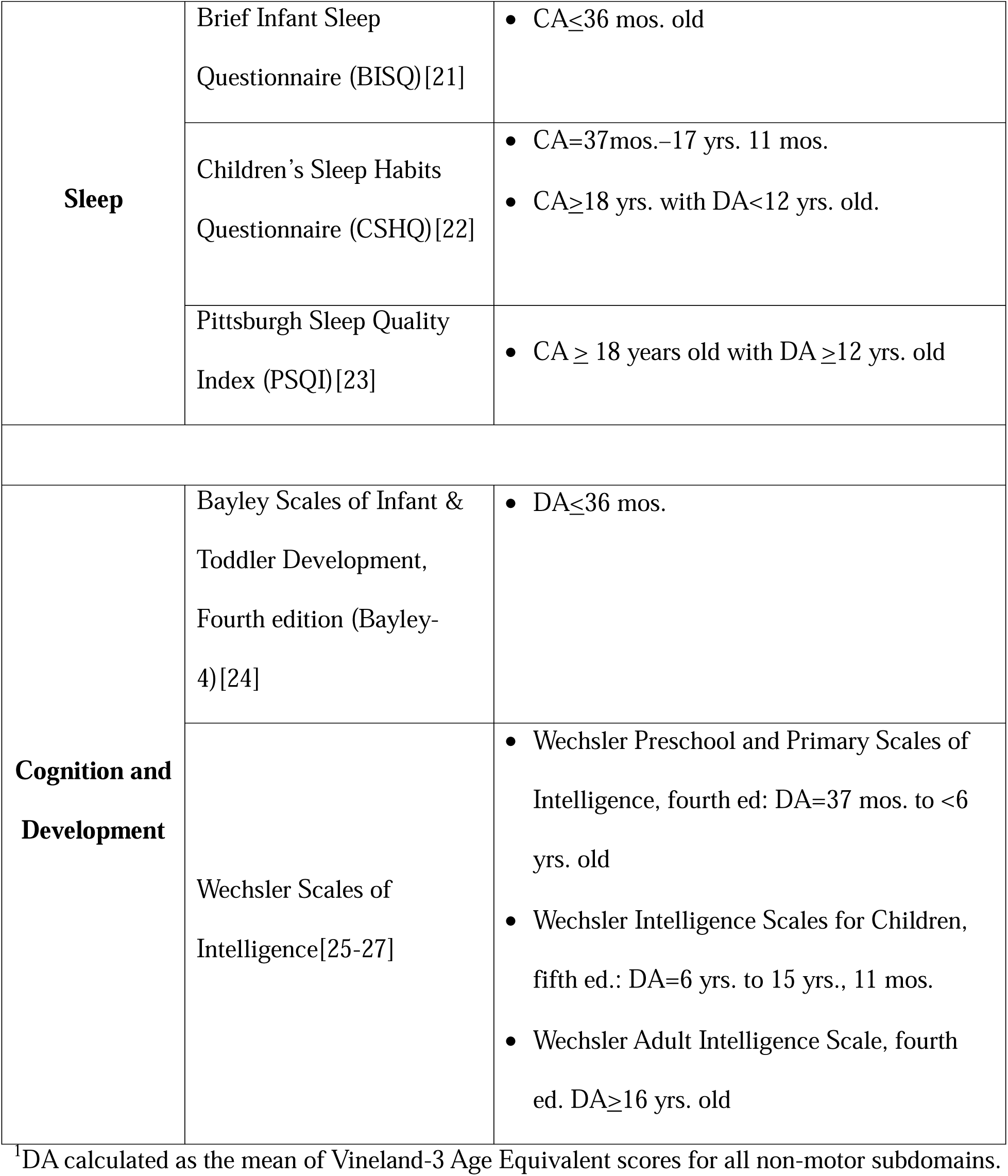
Measure Selection Based on Developmental Age (DA)^1^ or Chronological Age (CA)

### 2.4 Mood, Behavior, and Quality of Life

For most affected individuals, KCNT1-related epilepsy results in severe functional limitations. However, the range of phenotypic expression extends to milder forms affecting children and adults without severe encephalopathy, but with challenging behaviors, autism spectrum disorder, psychiatric symptoms, and/or functional impairments. For these affected individuals, we characterized such symptoms with a variety of assessments, described below. We also evaluated quality of life and adaptive skills for all affected individuals, including to determine whether existing tools to assess these domains were utile for this population.

#### 2.4.1 Child Behavior Checklist (CBCL)

The Child Behavior Checklist (CBCL) is an observer (parent)-reported measure to assess mood and behavior symptoms in school-age children [10]. The Preschool-age CBCL (P-CBCL ages 1.5 - 5 years old) also contains a Language Development Survey (LDS) to assess attainment of speech/language milestones and vocabulary acquisition [11]. The Adult Self Report form (ASR) and the Adult Behavior Checklist (ABCL; proxy report) were selected to assess adults with milder KCNT1-related epilepsy phenotypes [12].

#### 2.4.2 Screen for Child Anxiety Related Disorders (SCARED)

The SCARED is an observer (parent) – reported measure to assess recent anxiety symptoms and DSM-5 symptom diagnostic criteria for anxiety disorders in children ages 8-18 years old [13] and was used to assess children with an estimated developmental age of at least 8 years old based on the Vineland-3.

#### 2.4.3 Questionnaire for Psychotic Experiences (QPE)

In some adults with milder forms of KCNT1-related epilepsy, particularly the ADFNLE phenotype, psychiatric symptoms including hallucinations have been reported [14, 15]. The QPE is a self-report questionnaire to assess for the presence, severity, clinical features, and impact of delusional thoughts and hallucinations across various sensory modalities (e.g., auditory, visual, olfactory, etc.) and over two different time frames – recent (past week) and lifetime [16]. The QPE was used with any adult participants able to self-report their symptoms.

#### 2.4.4 Pediatric Quality of Life Inventory (PedsQL)

The PedsQL Infant and Core parent-proxy scales were used to assess health-related quality of life [17, 18]. Parents also completed the PedsQL Family Impact Module (FIM) to rate the impact of their child’s health condition on parental quality of life and general family well-being [19].

### 2.5 Sleep Quality

Disrupted sleep is a core component of sleep-related hypermotor epilepsy (SHE) in which focal seizures emerge during non-rapid eye movement (nREM) sleep stages [7, 20] and is also presumed to occur in other KCNT1 phenotypes due to the high daily burden of seizure activity. We selected several sleep questionnaires to assess various components of sleep function, including onset, duration, maintenance, quality, and parasomnias. The choice of instrument was guided by a participant’s chronological and developmental age.

#### 2.5.1 Brief Infant Sleep Questionnaire – Revised (BISQ-R) [21]

The BISQ is an observer (parent) reported questionnaire to assess an infant or toddler’s sleep patterns, including bedtime routine, sleep behaviors, daytime sleeping, and parental responses (e.g., how they help the child resume sleep after nighttime waking).

#### 2.5.2 Children’s Sleep Habits Questionnaire (CSHQ) [22]

The CSHQ is an observer (parent) reported measure. Domains assessed include: bedtime routines, asleep and awake times, sleep onset delay, sleep duration and persistence, sleep-disordered breathing, parasomnias, and sleep-related anxiety. Parents rated the presence, frequency, and impact of any sleep problems reported.

#### 2.5.3 Pittsburgh Sleep Quality Index (PSQI) [23]

The PSQI is a self-report measure. Items are grouped into subdomains capturing: sleep onset (latency), sleep quality, duration, persistence, and efficiency, sleep disturbances, daytime sleepiness, and use of sleep aid medications.

### 2.6 Physical measures

Head circumference (centimeters) was measured for Visits 1, 3, and 5 by the EEG technologist. At Visit 1, measurements were taken by a home study nurse for height, weight, BMI, temperature, heart rate, blood pressure, and respiratory rate. A 12-lead ECG was also performed.

### 2.7 Clinical Global impression (CGI) scale

The Clinical Global Impression Scale - Severity (CGI-S) score was a single, 7-point rating score of illness severity (1=normal, 7=severely ill) and was assigned by the investigator at Visits 1-5. The rating considered the participant’s current function and health status compared to the overall population of individuals with KCNT1 related epilepsy. At Visits 2-5, the CGI-Change (CGI-C) score was also assigned to describe change relative to a participant’s baseline assessment (1=much better, 7=much worse).

### 2.8 Laboratory Collections for Genetic and Biomarker Assessments

Noted above, plasma, CSF, serum and/or whole blood were collected for KCNT1 gene testing and biomarker research, including KCNT1 protein, proteins potentially associated with KCNT1, potential KCNT1 substrates, and biomarkers of neurologic disease including but not limited to neurofilament light chain (NfL) and neurofilament heavy chain (NfH). CSF was only donated (optional) from clinical samples; CSF biomarker research included evaluation of KCNT1 protein, proteins potentially associated with KCNT1, potential KCNT1 substrates, and NfL/NfH.

### 2.9 Cognitive and developmental assessments

To address a gap in the extant literature, we sought to systematically characterize cognitive, language, and motor milestones using normative-referenced, gold-standard assessments. Our additional objective was to assess the feasibility of using the selected measures with this patient population. The Vineland-3 mean AE score guided selection of developmentally suitable measures to assess participants, administered by a study Psychologist.

#### 2.9.1 Bayley Scales of Infant and Toddler Development, Fourth edition (Bayley-4) [24]

The Bayley-4 measures attainment of developmental milestones from infancy through 42 months of age. Because Bayley-4 assessment requires the child to engage with physical test materials (e.g., stacking blocks, pointing at pictures, etc.,), and involves motor tasks (e.g., grasping, crawling, etc.), it was only administered if participants opted to attend study visits in person at the clinical site.

#### 2.9.2 Wechsler Scales of Intelligence [25–27]

The Wechsler Scales of Intelligence measure intellectual ability (IQ). The specific test administered was guided by the participant’s developmental age (Table 1).

## 3. Sample Characteristics

35 participants (n=20 males, 15 females) enrolled and completed at least one assessment (i.e., baseline visit). The average age at the baseline visit was 76.0 months old (s.d. = 75.5), with most participants (n=28, 80%) at least 24 months of age. By contrast, developmental age was notably lower; the mean developmental age as estimated by the Vineland-3 was 8.5 months old (s.d. = 24.4). Most affected individuals were classified as having an EIMFS phenotype (n=29, 82.9%), with n=3 (8.6%) each having EOEE or SHE phenotypes. Table 2 presents detailed demographic characteristics of the sample.

**Table 2.** Demographic features of K1Te participants.

|  |  |
| --- | --- |
|  | **TOTAL (n = 35)** |
| **Chronologic age at screening (months)** |  |
| Mean (SD) | 76.0 (75.5) |
| Median | 53.0 |
| Q1, Q3 | 26.0, 112.0 |
| Min, Max | 1.0, 336.0 |
| < 24 months | 7 |
| ≥ 24 months | 28 |
| **Vineland-3 Developmental Age-Equivalent at screening (months)** |  |
| Mean (SD) | 8.5 (24.4) |
| Median | 0.00 |
| Q1, Q3 | 0.0, 12.0 |
| Min, Max | 0.0, 120.0 |
| **Sex, n (%)** |  |
| Male | 20 (57.1) |
| Female | 15 (42.9) |
| **Ethnicity, n (%)** |  |
| Hispanic or Latino | 2 (5.7) |
| Not Hispanic or Latino | 33 (94.3) |
| **Race, n (%)** |  |
| Asian | 1 (2.9) |
| Black or African American | 1 (2.9) |
| White | 30 (85.7) |
| Other / Mixed | 3 (8.6) |
| **Phenotype, n (%)** |  |
| EIMFS | 29 (82.9) |
| EOEE | 3 (8.6) |
| SHE | 3 (8.6) |
| **Duration of symptoms at screening, n (%)** |  |
| **≤ 2 years** | 8 (22.9) |
| EIMFS | 8 |
| EOEE | 0 |
| SHE | 0 |
| **> 2 years** | 27 (77.1) |
| EIMFS | 21 |
| EOEE | 3 |
| SHE | 3 |

| **Age at KCNT1-related epilepsy diagnosis (months)** |  |
| --- | --- |
| Mean (SD) | 34.9 (56.7) |
| Median | 0.0 |
| Q1, Q3 | 0.0, 60.0 |
| Min, Max | 0.0, 240.0 |

## 4. Safety considerations

Participants and their parents were monitored for any study-related risks. These included potential for complications to participants from blood draws, EEG-related risks, completion of neurobehavioral assessments and questionnaires, and risks to privacy and confidentiality. Only qualified vendors were used for home-based study procedures.

Because seizure frequency and type were primary outcomes, they were not considered adverse events, except for status epilepticus, a clinically significant increase in seizure frequency or intensity, or emergence of a new seizure type. During the EEG, a remote technologist monitored the video feed and EEG recordings; if a seizure lasted more than 3 minutes, the technologist could call the parent or local emergency number if needed. Also, parents were advised to remain with the child for the duration of the EEG, i.e., sleeping in the same room.

## 5. Discussion

### 5.1 Fully-decentralized operations in a severe developmental and epileptic encephalopathies during the COVID-19 pandemic

The initial protocol design offered participants the option of conducting study visits remotely or in-person at the clinical site. During the study design phase, patient advocacy and community feedback strongly suggested that the overwhelming majority of visits, if not all, would be opted to be done remotely, due to the potential health complications and parental challenges associated with participant travel. Additionally, enrollment and study visits were conducted from August 2021 to August 2023, with ebbs and rises in community rates of SARS-CoV-2 (COVID-19) during this time. Therefore, the study converted to fully-remote, home-based operations. Nonetheless, this introduced potential risk of exposure to COVID-19 and other communicable illnesses via in-home research staff. A safety protocol was established including rescheduling home visits if visiting staff or family members were unwell, and to notify family members of any potential exposures, i.e., if staff visiting the home subsequently tested positive for COVID-19. Additionally, before completion of home-based EEG, a health screening questionnaire was completed to document whether the participant or any household member had signs or symptoms of, or exposure to someone with any upper respiratory illnesses, and a known history of allergy to adhesives or latex.

### 5.2 The role of technology in the home-based research visit

Due to the need for high-quality, contextually relevant prospective and longitudinal data to advance clinical care and research, substantial efforts were made to accomplish all study activities within the home at the level of rigor expected of a clinical trial. This required leveraging certain technologies. An internet-capable tablet was used to perform electronic consent as aided by a virtual telehealth visit or phone call, collect seizure endpoints via an electronic diary, and administer patient- or parent-reported outcomes. Good Clinical Practice (GCP) standards were used when available. Telemedicine visits were completed on parents’ personal internet-enabled devices using a HIPAA-compliant Zoom video conferencing platform (Zoom Video Communications, Inc.).

### 5.3 Video EEG

Video EEG monitoring is the gold standard to evaluate for epileptic activity. Ambulatory EEGs without video are suboptimal in this patient population who typically have very abnormal EEG backgrounds and movements that are not necessarily ictal. Video EEG monitoring within an epilepsy monitoring unit is ideal, but difficult for families due to travel burden and/or other childcare responsibilities, and places the child at risk for nosocomial infections. At-home, prolonged video EEG allowed for longer EEG recordings while minimizing burden and risk.

### 5.4 Challenges and Learnings

#### 5.4.1 Home-based EEG

Home-based overnight video EEGs worked well in most households. Parents received guidelines to ensure the EEG video capture was adequate, e.g., ensuring that participants remained in view of the camera and were not overly obscured by blankets throughout the monitoring period. Most participants were non-ambulatory had few independent movements, which facilitated stable placement of EEG leads. However, parents were given additional materials and instructions for re-attaching leads if disconnected. Secure upload, storage, and review processes were established between the site and vendors.

#### 5.4.2 Lab draws

Home-based collection of blood samples in developmentally delayed participants can be challenging due to the absence of hospital-based resources to support collection of labs in participants with difficult-to-access vasculature. The protocol allowed one reattempt on a separate date, but if that also failed, then no sample was collected for that participant.

#### 5.4.3 Technology literacy

Parents had varying levels of literacy and comfort with the technologies required for study participation, particularly the use of the tablet device to complete questionnaires and the electronic seizure diary. The Coordinator met with parents via a secure online video meeting soon after enrollment to provide remote training, and assisted with troubleshooting throughout the study. The Coordinator also received direct, ad hoc technical assistance from the vendor who programmed and supplied the tablets.

#### 5.4.5 Visit windows

The study protocol included multiple types of home-based activities reliant upon different healthcare vendors (EEG, ECG, phlebotomy, etc.) deployed to participant homes across the United States. Flexibility for scheduling within designated multi-day visit windows was appreciated by participating families and vendors alike. Consistent with this population, some participants required hospitalization for illness or procedure, necessitating rescheduling of study activities. A 14-day study visit window, and a 21-day window for the baseline visit (with the most activities to coordinate), provided flexibility to complete all study activities while minimizing protocol deviations.

#### 5.4.6 Cognitive and developmental assessments

With a fully-remote study, some in-person assessments (e.g., Bayley-4) could not be completed. Future studies of KCNT1-related epilepsy should evaluate feasibility and utility of these assessments, either with participants attending visits at the study site, or possibly through “research house calls,” i.e., study personnel conducting assessments at participants’ homes. The latter approach requires an adequately controlled home environment (i.e., quiet, non-distracting, private) that emulates the conditions for research-grade cognitive and developmental evaluations, and should consider operational challenges of specialists covering large geographic territories.

#### 5.4.7 Wearable devices

Behavioral barriers may limit a participant’s ability to tolerate wearable devices such as a wrist-worn monitor. It can be helpful for the study team to provide guidance to parents for a gradual and positive introduction to these items (e.g., encouraging participants to first touch/handle the item, then wear for increasing amounts of time) and to associate use of wearable devices with preferred activities.

#### 5.4.8 Study adherence

Remote endpoints require participants to complete study activities on schedule in an unsupervised environment. Reminders may prompt the participant or parent to keep a device charged and connected to the internet, receive and respond to communications from study personnel, and/or attend telehealth visits. The study team maintained ongoing communication with parents to provide encouragement, manage adherence for remote assessments, and troubleshoot issues for those struggling to maintain adherence. Some non-adherence was anticipated due to medical issues of the affected individual that could detract from parents’ time and attention to study activities. The remote study design helped mitigate this burden by providing greater flexibility for study activity scheduling and removing the travel burden. Future studies should also anticipate and support patients and their parents who may experience these often unavoidable, and disease-related barriers to adherence.

## 6. Conclusion

The K1Te study allowed for prospective and longitudinal measurement of the natural history of of KCNT1-related epilepsy using a fully-decentralized design. This study design captured participant and parental burden of disease while minimizing demands of study participation, in a population for whom in-person study visits are challenging and may influence outcome measures. Results and data analysis will be separately reported when available. Future research efforts can consider these design choices, and challenges and learnings, to develop better therapies and care decisions for individuals impacted by KCNT1-related epilepsy and other rare developmental and epileptic encephalopathies.

## Glossary

ADNFLE: Autosomal Dominant Nocturnal Frontal Epilepsy;
EIMFS: epilepsy of infancy with migrating focal seizures;
EOEE: early-onset epileptic encephalopathies;
K1Te: A Natural History Study of Participants with Potassium-Sodium-Activated Channel Subfamily T Member 1 (KCNT1)-Related Epilepsy;
KCNT1: Potassium-Sodium-Activated Channel Subfamily T Member 1;
SHE: sleep-related hypermotor epilepsy

## Acknowledgments

This research was supported by a grant to the University of Rochester from Biogen. We thank the affected individuals and parents who participated in the research. For assistance with recruitment, we thank the KCNT1 Epilepsy Foundation.

## Author Contributions - CRediT

### Author contributions

**Heather Adams:** Conceptualization, Methodology, Original draft preparation, Review & Editing; **Viet Nguyen:** Conceptualization, Methodology, Review & Editing; **Laurie Seltzer:** Review & Editing; **Carolyn Dickinson:** Review & Editing; **Courtney Aponte:** Review & Editing; **Sarah Hubbard:** Conceptualization, Methodology, Review & Editing; **Marco Rizzo:** Conceptualization; Methodology, Review & Editing; **David Bearden:** Conceptualization, Methodology, Original draft preparation, Review & Editing;

## Funding sources

Study Sponsor: This work was supported by a research grant from Biogen

## Data availability

Data sharing not applicable as no datasets were generated or analyzed for this methods paper.

## Ethics Approval

All research activities were conducted under an IRB-approved protocol (WCG IRB Tracking Number 20212955)

## Participant Consent

Parents provided written permission for participation of their affected child/family member. Due to the severe cognitive impairment associated with KCNT1-related epilepsy, the IRB granted a waiver of assent for affected children and waived consent for affected adults.

## Notes

### Competing Interest Statement

Heather Adams received salary support to conduct this research through a grant from Biogen and previously served as a paid consultant to Biogen.
Viet Nguyen received support from Biogen in connection with this work as an employee.
Sarah Hubbard received support from Biogen in connection with this work as an employee.
Marco Rizzo received support from Biogen in connection with this work as an employee and owns Biogen stock.
Laurie Seltzer received salary support to conduct this research through a grant from Biogen.
Carolyn Dickinson received salary support to conduct this research through a grant from Biogen.
Courtney Aponte received salary support to conduct this research through a grant from Biogen.
David Bearden received salary support to conduct this research through a grant from Biogen. He has served as a paid consultant for Biogen, Servier, Actio Bio, and UCB. Dr. Bearden serves in an upaid role as the Chairperson of the Scientific Advisory Board for the KCNT1 Foundation.

### Author Declarations

IRB of WCG gave ethical approval for this work (WCG IRB Tracking Number 20212955)

